# Non-Adherence Tree Analysis (NATA) - an adherence improvement framework: a COVID-19 case study

**DOI:** 10.1101/2020.06.30.20135343

**Authors:** Ernest Edifor, Regina Brown, Paul Smith, Rick Kossik

**Affiliations:** Operations, Technology, Events and Hospitality Management, Manchester Metropolitan University, Manchester, Lancashire, UK; Medicine, University of Massachusetts Medical School, Worcester, Massachusetts, USA; Marketing, Retail and Tourism, Manchester Metropolitan University, Manchester, Lancashire, UK; Research and Development, GoldSim Technology Group LLC, Seattle, Washington, USA

**Keywords:** non-adherence, fault tree analysis, Monte Carlo simulation, COVID-19

## Abstract

Poor adherence to medication is a global phenomenon that has received a significant amount of research attention yet remains largely unsolved. Medication non-adherence can blur drug efficacy results in clinical trials, lead to substantial financial losses, increase the risk of relapse and hospitalisation, or lead to death. The most common methods measuring adherence are post-treatment measures; that is, adherence is usually measured after the treatment has begun. What the authors are proposing in this multidisciplinary study is a technique for analysing the factors that can cause non-adherence before or during medication treatment.

Fault Tree Analysis (FTA), allows system analysts to determine how combinations of simple faults of a system can propagate to cause a total system failure. Monte Carlo simulation is a mathematical algorithm that depends heavily on repeated random sampling to predict the behaviour of a system. In this study, the authors propose the use of Non-Adherence Tree Analysis (NATA), based on the FTA and Monte Carlo simulation techniques, to improve adherence. Firstly, the non-adherence factors of a medication treatment lifecycle are translated into what is referred to as a Non-Adherence Tree (NAT). Secondly, the NAT is coded into a format that is translated into the GoldSim software for performing dynamic system modelling and analysis using Monte Carlo. Finally, the GoldSim model is simulated and analysed to predict the behaviour of the NAT.

This study produces a framework for improving adherence by analysing social and non-social adherence barriers. The results reveal that the biggest factor that could contribute to non-adherence to a COVID-19 treatment is a therapy-related factor (the side effects of the medication). This is closely followed by a condition-related factor (asymptomatic nature of the disease) then patient-related factors (forgetfulness and other causes). With this information, clinicians can implement relevant measures and allocate resources appropriately to minimise non-adherence.

## Introduction

A great proportion of patients (especially those with chronic diseases) are non-adherent to their medication regimen [1,2]. This has led many researchers to the conclusion that non-adherence poses a significant challenge in medical practice [3,4]. Some authors [5] class non-adherence as an “epidemic”, while the World Health Organisation (WHO) [1] considers it as “a worldwide problem with striking magnitude”. Patients’ non-adherence to treatment interventions could have grave consequences; it could blur the efficacy of treatments [6], create large financial costs to sponsors [7], cause adverse events or even lead to death in some cases [8].

Non-adherence to medications is not limited to any particular disease – acute or chronic; it affects all diseases [9] and can be influenced by the timing, consistency and persistence of taking medications. Barriers to medication adherence can vary significantly, ranging from patient-related barriers to treatment-related barriers. Care providers, the healthcare system and medical staff also contribute to non-adherence [4,10]. Given this variation in barriers to adherence, there is no single intervention that will effectively minimise medication non-adherence [4,11]. For example, behavioural modification is one way to improve adherence however, this is a very challenging solution to implement as human behaviour is not easily altered. Behavioural modification can take the forms of education, motivation, support and monitoring [12]. Tackling individual aspects of non-adherence can be done, however, there is a need for a multidisciplinary approach to medication non-adherence [12].

There are various techniques for assessing non-adherence. Though some are classic, such as pill counting, others employ more sophisticated approaches [13]. Methods for measuring medication adherence can be generally put in two main categories: direct and indirect [3]. The former provides proof that patients have taken their medication as prescribed while the latter cannot provide such proof. Direct methods include body fluid sampling, direct observation of patient and measurement of biological markers [6]. Indirect methods, which are more widely implemented, include pill count, patient questionnaire [14], self-report forms, and electronic monitoring devices. Medication adherence is characterised by three main components: initiation (the point when the patient takes the first dose as prescribed), implementation (period of dosing regimen complying with prescription) and discontinuation (the point when the patient stops taking medication as prescribed) [15].

Measuring medication adherence can be challenging due to the use of adherence measures that have poor accuracy and reliability [16]. Most of the methods for measuring adherence are performed during the implementation phase of adherence [3]. Sometimes adherence measurements are performed during the discontinuation phase [17]. There is limited literature on the methods for measuring adherence before the initiation phase. Self-report methods of measuring adherence are usually performed during the implementation phase. However, these self-reporting tools can be used as historical data to measure adherence before the initiation phase of other future treatments. The Medication Adherence Reasons Scale (MAR-Scale) and the Morisky Medication Adherence Scale (MMAS) can be used to measure adherence before the initiation stage [18,19]. Knowing the common reasons for a patient’s non-adherence to medications that they take for their chronic medical conditions can help clinicians or pharmacists design interventions that will increase the chances of the patient adhering to the new medication before the patient starting the medication. The MMAS scale requires the patient to have other chronic medical conditions for which they are taking medications. The MAR-Scale is unable to fully capture and analyse system conditions that may contribute to non-adherence but may not be directly associated with the patient or the medication.

Various techniques proposed for improving adherence are complex and ineffective, therefore, they are unable to realise the full benefits a treatment could deliver [16]. It is rational to assess and measure patients’ likely non-adherence before the initiation stage of medication treatment to improve adherence. The authors employ a proven probabilistic risk assessment technique to estimate the likelihood of non-adherence before the initiation stage. The results of this study help clinicians to identify and assess barriers to adherence; this aids them in the development of non-adherence mitigating strategies and allocation of resources to improve adherence before the initiation stage of medication adherence.

## Method

### Fault Tree Analysis (FTA)

Fault Tree Analysis (FTA) [20], since its inception, has been used mostly in the engineering sector. It is a tree-like graphical representation of how basic components failures (basic events) of a system can propagate to cause a total system failure (top-event). Events are logically connected using the Boolean gates AND, OR and sometimes the Priority-AND (PAND) gates – depicted in Fig 1. The AND gate (conjunction) represents the situation where all children events of an output event need to occur for the output event to occur. The OR (disjunction) gate represents the situation where at least one child event of an output event need to occur to trigger the occurrence of the output event. The Priority-AND (PAND) gate represents the scenario where all the children events of an output event occur in a strict sequence – one after another – for the output event to occur.

**Fig 1.**
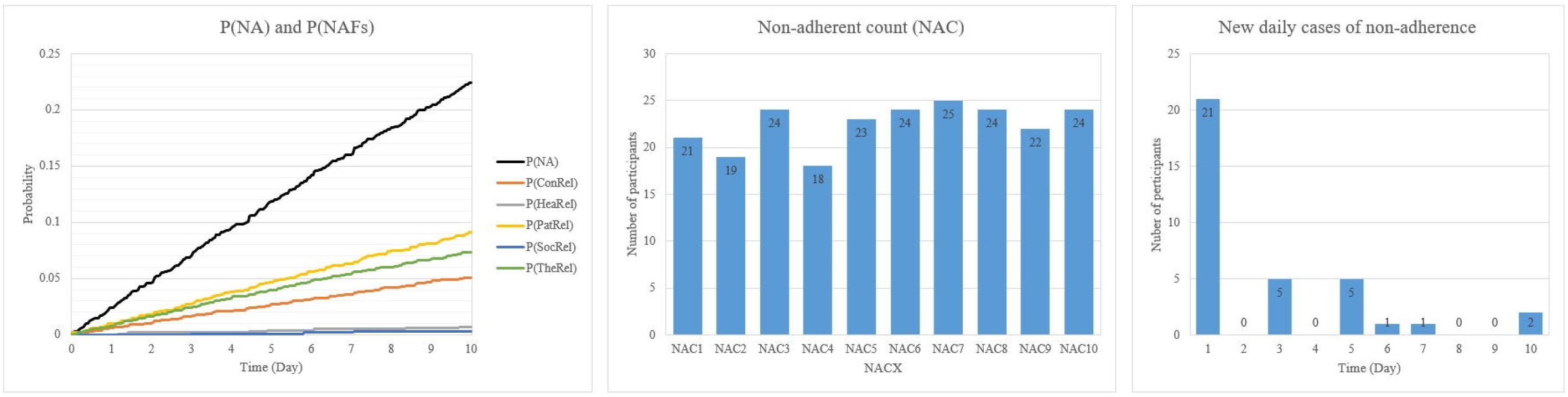
FTA Logic Gates.

In Fig 1X, the output event *Q* is triggered when its input (or child) events *A* and *B* have occurred within a given time, *t*. In Fig 1Y, the output event *Q* is triggered after a given time, *t*, when at least one of its input events – *A* or *B* – have occurred. Fig 1Z depicts the PAND gate where the output event *Q* is triggered after a given time *t* only when its input events *A* occurs before *B*. For a detailed description of how FTA is performed, the reader is referred to Vesely et al. [20]. In a logical expression, the AND, OR and PAND gates are represented by the symbols, *, + and < respectively.

Once a system has been translated into a fault tree, it can be analysed logically (qualitatively). The logical analysis involves the determination of minimal cut sets (MCS) using Boolean algebra. MCS is the smallest combination of basic events that are *necessary* and *sufficient* to cause the top event. *Necessary* means each basic event in the MCS is needed for the top event to occur and *sufficient* means the MCS does not need the occurrence of additional events to cause the top event occurrence. In addition to creating MCS, the logical analysis also reveals single points of failure of a system and reveals relationships between components. Quantitative analysis or probabilistic analysis involves the evaluation of the probability of the system failing using the MCSs. The probability, *P*, of the PAND, AND and OR (in order of precedence), within a given time *t*, for an events *X*_*1*_…*X*_*n*_ can be calculated using Equations (1), (2) and (3) respectively [20]. Equation (1) is limited to exponentially distributed independent events.

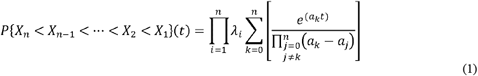

Where *a*_*0*_ = 0 and 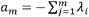 for *m* > 0.

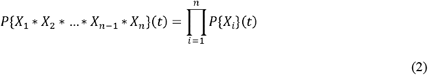

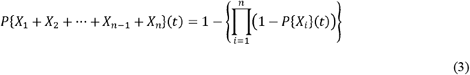

To improve the overall reliability of the system, one could perform criticality/sensitivity analysis [20] to determine how individual components contribute to the system failure. The results of a sensitivity analysis enable investigators to implement mitigating strategies, know the quality of components to use and allocate resources appropriately to improve the overall reliability of the system.

### GoldSim Software

Most traditional FTA-based techniques have some limitations; they are mostly limited to analytical approaches with exponentially distributed component failures, they cannot capture repairable events and they are unable to process other system environment data, such as the time of operation. To evaluate real-world scenarios, one needs to overcome such limitations because real-world events are dynamic, mostly repairable, and could have different failure distributions. These limitations are addressed by GoldSim software [21]. GoldSim is a software capable of performing the modelling and probabilistic analysis of complex real-world systems using Monte Carlo simulation. It has features for representing the classical Boolean gates AND and OR. The PAND gate can be modelled accurately using dynamic and intuitive elements in GoldSim.

### Non-Adherence Tree Analysis (NATA)

The authors propose the Non-Adherence Tree Analysis (NATA) – a systematic and holistic technique heavily based on the fault tree analysis technique. Unlike FTA where the primary focus of the investigation is the reliability of a system using failure data (such as failure rate), the primary focus on the investigation in NATA is non-adherence (or discontinuation of medication) using factors that trigger non-adherence. NATA follows the guidelines of the classical FTA, however, new terms are defined in this study to reflect its novel domain of application; these terms have been adapted from classical FTA definitions [20]. Examples will be based on this scenario: *in a study, 20 patients (out of 100 patients) fail to take their medication as prescribed during a 10-day medication regimen. Out of the 20 non-adherent patients, 6 were non-adherent due to forgetfulness (FORG), 4 due to side effects (SIDE) and 10 due to other factors (OTHER)*.

#### Days of Medication Adherence (DoM)

This is the total number of days a medication should be taken in a treatment regimen for a given study.

#### Number of participants (NoP)

This is the total number of participants in a study.

#### Non-Adherence (NA)

This is known as the top event in classical FTA. NA represents the situation where a prescription to a medication regimen has not been followed as instructed. Meaning, NA is discontinuation in adherence to the medication before the end of prescribing period [15].

#### Non-adherence tree (NAT)

A graphical top-down deductive structure that represents the non-adherence factors as nodes with Boolean logic gates connecting these nodes to show the relationship between them. Fig 2 is a simple NAT for the scenario. When creating NATs, additional information such as the time of operation, replacements, repair/resolution, etc., can be included in the rectangle of the corresponding non-adherence factor.

**Fig 2.**
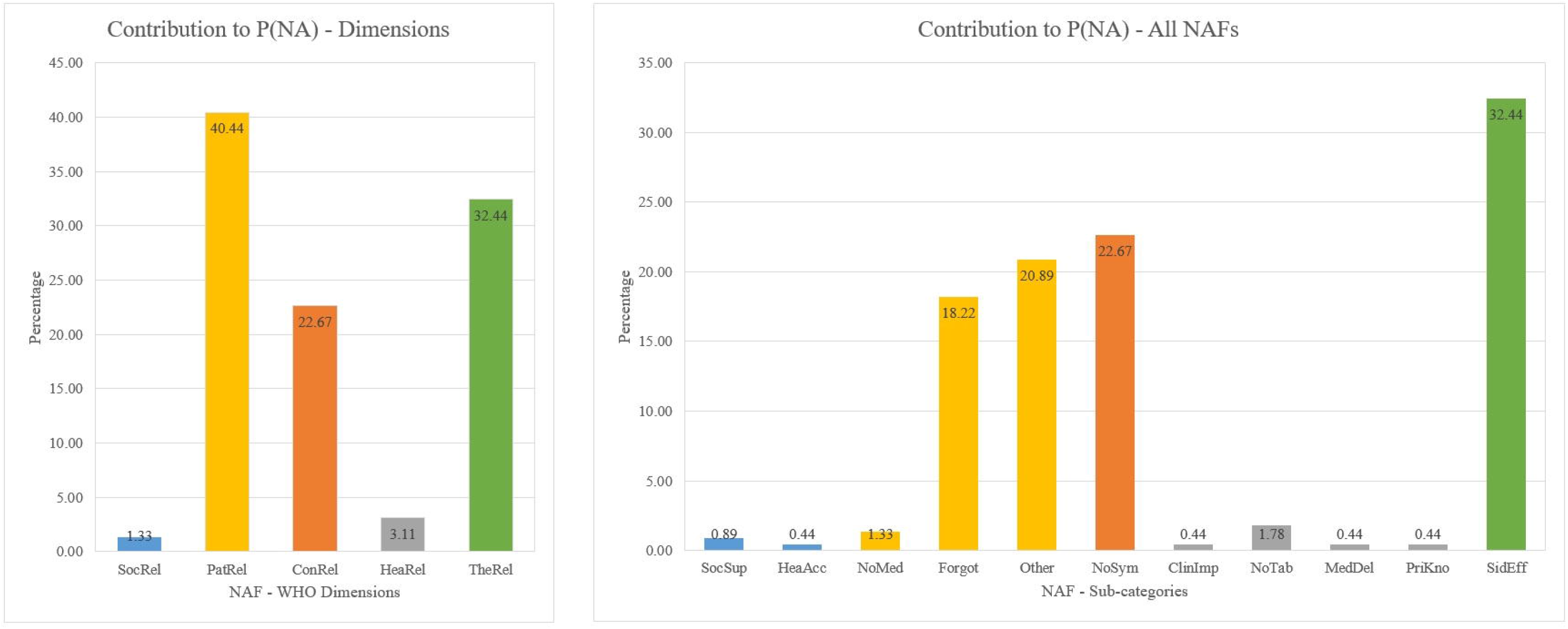
A Non-Adherence Tree (NAT)

#### Non-Adherence Factor (NAF)

This is synonymous to an event in FTA. It is a binary outcome indicating if a factor leading to medication discontinuation has occurred or not. It is true when the factor has occurred and false otherwise. This could be *FORG, SIDE* or *OTHER*.

#### Basic NAF

This is synonymous to a basic event in FTA. It is a discrete NAF that cannot be decomposed into other NAF. This is *FORG* and *SIDE*; *OTHER* could be broken into other NAFs if need be.

#### Non-Adherence Count (NAC)

This is the cumulative number of patients who are non-adherent in a study, clinical trial or medication administration process. This is calculated as the sum of the count of all the occurrences of NAFs. This can be expressed as:

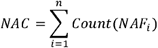

From the scenario,

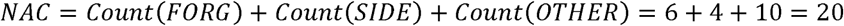

NAC could also be expressed in terms of the number of days elapsed using NAC*X*, where *X* is the number of days. For example, NAC3 = 8, means 8 participants were non-adherent by Day 3 (counting from Day 1) of a medication regimen.

#### GrandNoP

this represents the total number of non-adherent patients from *n* different studies and it can be evaluated as:

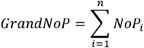

#### Non-Adherent Rate per Study (NARS)

This is the number of occurrences of a particular NAF per the NoP and can be defined as:

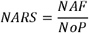

Therefore, the NARS for *FORG* can be evaluated as

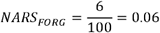

#### Non-Adherent Rate (NAR)

This is synonymous to failure rate or hazard function in FTA. NAR is the rate of occurrences (NAR) per the duration of the medication regimen – usually expressed in days. NAR can be expressed as,

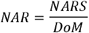

Therefore, the NAR for *FORG* can be evaluated as

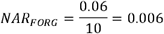

#### Weighted NAR (WNAR)

where NARs are to be sourced from multiple studies, it is useful to have a weighted NAR. The WNAR for a particular NAF from *n* studies can be evaluated

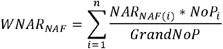

#### Non-Adherence Factor Probability

This is the probability that a particular NAF will occur and it is represented by P(NAF). The determination of the P(NAF) is based on the probability distribution of the NAF. For example, given a duration (*d*) of 1 and 10 days respectively, if *FORG* is exponentially distributed, P(*FORG*) can be evaluated as:

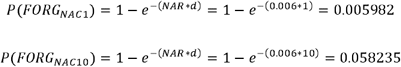

#### Non-Adherence Probability

This is the overall non-adherence probability – the probability that there will be discontinuation as a result of NAFs at the end of a medication regimen.

Represented by P(NA), the non-adherence probability can be evaluated from Eqn 3. Therefore, from the scenario, given a duration (*d*) of 10 days, P(NA) is

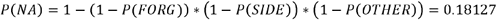

## Results

To demonstrate the usefulness of NATA, it is applied to a COVID-19 treatment intervention clinical trial. Several clinical trials of drugs targeting COVID-19 have been registered in China [22]. Remdesivir, a nucleotide analogue, and chloroquine, an anti-malarial compound, have both shown inhibition of the new coronavirus [23]. As of April 2020, several clinical trials are testing the therapeutic efficacy of remdesivir and hydroxychloroquine (ClinicalTrials.gov: NCT04280705, NCT04329923) for COVID-19 treatment. If any of these drugs are shown to be safe and efficacious, they could become the first drug approved for the treatment of COVID-19. In a hospitalized setting, “there is less consideration given to adherence” [24] therefore, this study will only consider the out-patient settings. It is assumed that the treatment for out-patients, who usually have mild symptoms, is a tablet that will be administered for 10 days by the patients themselves - one pill per day - for a study population of 1000 patients. The diagram in Fig 3 is a NAT for a hypothetical treatment intervention for COVID-19 using the WHO’s dimensions [1,5] and NAFs from six studies [25–30].

**Fig 3.**
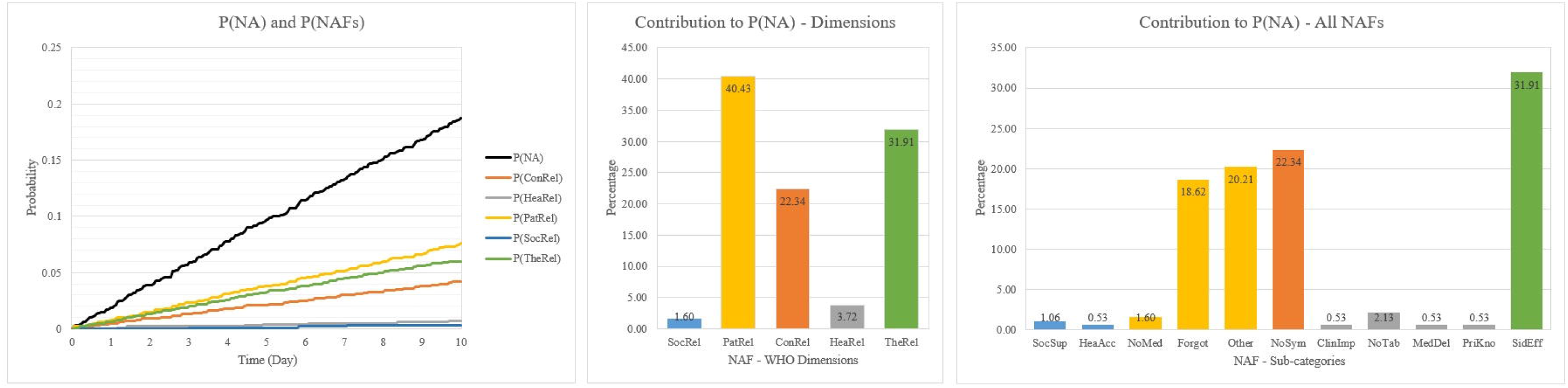
A NAT for COVID-19 Intervention.

In Fig 3, non-adherence has been classified into the 5 WHO dimensions: Social/Economic-related factors (*SocRel*), Patient-related factors (*PatRel*), Condition-related factors (*ConRel*), Healthcare-related factors (*HeaRel*) and Therapy-related factors (*TheRel*). These top-level NAFs have sub-NAFs that are based on factors for non-adherence of oseltamivir, an oral antiviral medication that inhibits influenza viral replication. This antiviral medication is chosen because of the similarities in symptoms between influenza infection and COVID-19. Six studies (covering different demographics and geographic locations) have been used in the determination of these sub-NAFs; they are henceforth referred to as *Study 1* [25], *Study 2* [26], *Study 3* [27], *Study 4* [28], *Study 5* [29] and *Study 6* [30].

In general, there is a strong correlation between family/social support networks for patients and their adherence to a medication regimen [31]. Patients with COVID-19 require self-isolation to avoid the spread of the disease. Therefore, limited social support (*SocSup*) and limited healthcare access (*HeaAcc*) have been considered as NAFs contributing to *SocRel*. Since this intervention is novel, it is assumed that a NAF in the *HeaRel* category is “lack of prior knowledge of adherence” (*PriKno*) [1,5] in addition to limited tablets (*NoTab*) and clinical improvement (*ClinImp*). The medicine delivery system in the hospital can also contribute to *HeaRel* if both the ICT (*IctSys*) and manual (*ManSys*) delivery systems fail. NAFs contributing to *PatRel* include patients’ forgetfulness (*Forgot*), choice of not taking the medication (*NoMed*) and other patient-related factors (*Other*). *ConRel* and *TheRel* have only one NAF each – no symptoms (*NoSym*) and side effects (*SidEff*) respectively.

## Logical Analysis

Using basic Boolean logic, the MCS for non-adherence can be evaluated as:

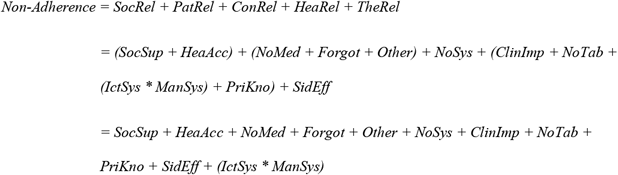

The MCS reveals that there are ten single points of failure in the system. Both *IctSys* and *ManSys* need to occur together to trigger discontinuation therefore they are not considered single points of failure. With a quick scan at these single points of failure, investigators can determine which aspect of the system need backups. For example, *NoTab* is a factor that could be easily and quickly improved to enhance adherence; not all the other factors can be quickly improved. For a detailed analysis on which factor contributes most to non-adherence, probabilistic analysis is required. Probabilistic analysis can only occur when NARs have been determined.

## Non-Adherence Rates (NARs)

It is assumed that the recruited ambulatory participants would fail to adhere to their medication due to *HeaAcc* resulting in a WNAR of 1.2E-4/day. It is also assumed that *PriKno* [1,5], has an initial WNAR of 1.5E-4. This rate reduces by 8 per cent of the initial WNAR multiplied by the number of elapsed day to represent the increasing knowledge of adherence by the medical team. The *IctSys* and *ManSys* sub-systems responsible for ordering and dispensing the medicine fail at daily rates of 8.12E-5 and 5.34E-5 with mean-delay-time-until-repair of 4 hours and 2 hours respectively. There is a 2-day delay time until the medication is delivered in case of *NoTab*. From results presented in Belmaker et al. [25], it is estimated that the WNAR (per day) for *SocSup* for patients taking oseltamivir who are under age 25, between ages 25 and 45 inclusive and over age 45 are 4.138E-4, 1.379E-4, and 2.069E-4 respectively. Table 1 is a summary of NAFs, NARSs, NARs and WNARs from the six studies [25–30].

**Table 1.** NAFs, NARSs, NARs and WNARs from six studies.

| Study (S) | NoTabs | SidEff | NoMed | ClinImp | Forgot | NoSym | Other |
| --- | --- | --- | --- | --- | --- | --- | --- |
| **S1 (NoP=201, DoM=10)** | 5 | 3 | 4 | - | - | - | 13 |
| NARS | 2.488E-02 | 1.493E-02 | 1.990E-02 | - | - | - | 6.468E-02 |
| NAR (per day) | 2.488E-03 | 1.493E-03 | 1.990E-03 | - | - | - | 6.468E-03 |
| **S2 (NoP=33, DoM=5)** | - | 1 | - | 1 | - | - | 4 |
| NARS | - | 3.030E-02 | - | 3.030E-02 | - | - | 1.212E-01 |
| NAR (per day) | - | 6.061E-03 | - | 6.061E-03 | - | - | 2.424E-02 |
| **S3 (NoP=331, DoM=6)** | - | 9 | - | - | 21 | - | 7 |
| NARS | - | 2.719E-02 | - | - | 6.344E-02 | - | 2.115E-02 |
| NAR (per day) | - | 4.532E-03 | - | - | 1.057E-02 | - | 3.525E-03 |
| **S4 (NoP=313, DoM=7)** | - | 20 | - | - | - | 42 | 16 |
| NARS | - | 6.390E-02 | - | - | - | 1.342E-01 | 5.112E-02 |
| NAR (per day) | - | 9.128E-03 | - | - | - | 1.917E-02 | 7.303E-03 |
| **S5 (NoP=326, DoM=5)** | - | 24 | - | - | 7 | 13 | 4 |
| NARS | - | 7.362E-02 | - | - | 2.147E-02 | 3.988E-02 | 1.227E-02 |
| NAR (per day) | - | 1.472E-02 | - | - | 4.294E-03 | 7.975E-03 | 2.454E-03 |
| **S6 (NoP=246, DoM=10)** | - | 24 | 1 | - | 22 | - | 9 |
| NARS | - | 9.756E-02 | 4.065E-03 | - | 8.943E-02 | - | 3.659E-02 |
| NAR (per day) | - | 9.756E-03 | 4.065E-04 | - | 8.943E-03 | - | 3.659E-03 |
| **WNAR** | 3.448E-04 | 8.315E-03 | 3.448E-04 | 1.379E-04 | 4.897E-03 | 5.931E-03 | 5.002E-03 |

## Probabilistic Analysis

A GoldSim model was created from the NAT in Fig 3 using the data in Table 1. The system was simulated over 10 days with a time-step of one hour. For each time-step, 1000 iterations were performed to simulate the behaviour of each participant. Fig 4 is a graph depicting the mean P(NA) and the NAC over ten days. P(NA) on Day 0 is zero, however, as the days progress towards Day 10, it approaches 1; on Day 10, it reaches 0.22 with a 5% and 95% confidence bounds of 0.2 and 0.25 respectively and a standard deviation of 0.42. Missing at least one pill (of the 10 pills) results in non-adherence. The results predict that 776 participants would take all their medications (10 pills) as prescribed in ten days; 224 participants would miss at least one pill. This result aligns with the results of the six studies: adherence to such treatment is very high. The NAF contributing the most to P(NA) is the *PatRel*. Meaning, patient-related factors are strongly correlated to non-adherence.

**Fig 4.**
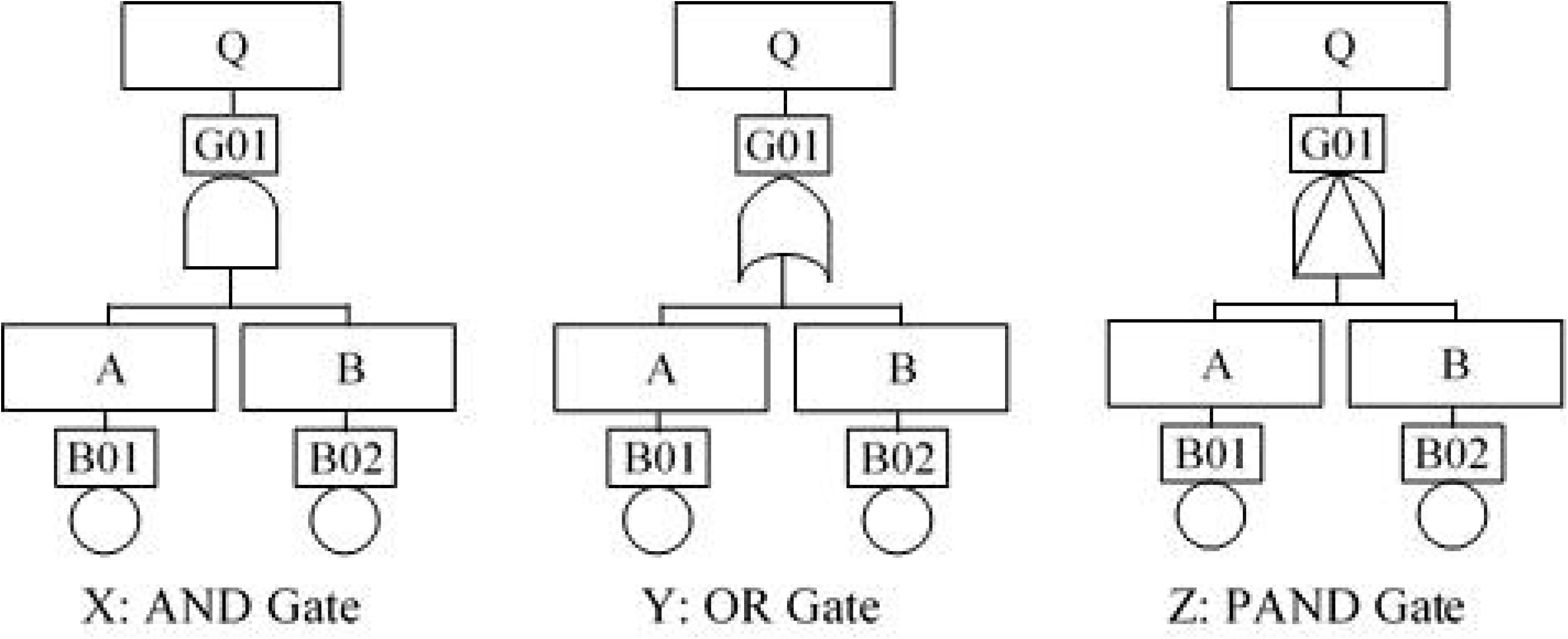
Probability of Non-Adherence and Non-Adherent Participants.

Cumulatively over the 10 days, the average number of patients who would be non-adherent increases steadily. By the end of Day 1, it is estimated that 21 would miss their pills. From Day 1 to the end of Day 5, it is estimated that 105 participants would miss at least one pill. At the end of the medication regimen, it is estimated that 224 patients would miss at least one of their pills. However, the number of new daily cases on non-adherence had no clear pattern; there were no new cases on some days whiles only 2 new cases of non-adherence would be observed on Day 10. The average NAC to NoP ratio in the six studies is 0.17; the NAC to NoP ratio for the COVID-19 case study is 0.22. The difference in ratios is due to the additional NAFs (such as *SocSup, HeaAcc* and *PriKno*) added to give a more accurate dynamic of the behaviour of non-adherence to the COVID-19 disease.

## Discussion

Based on the NATA results for the case study, a COVID-19 treatment is likely to have the non-adherence probability predicted in Fig 4. Given the high rate of contagiousness, significant financial and economic burden, and the number of deaths COVID-19 has caused, there is a need for the result to improve – that is, increase overall adherence rate. At a glance, it seems that patient-related factors contribute the most to non-adherence. However, patient-related factors are not solely responsible for non-adherence; other factors also contribute to non-adherence – this affirms results in previous studies [1]. Further investigation of the results gives us a different picture. In Fig 5, it can be seen that patient-related factors contribute about 40% to the non-adherence probability. However, when the constituent NAFs are considered, *SidEff* is the biggest contributor, closely followed by *Forgot, NoSym, Other*, etc. This means that clinicians hoping to improve the patients’ adherence to a COVID-19 treatment should concentrate on reducing these factors – more importantly, the *SidEff. NoSym* is known to have a relatively high ratio in known COVID-19 cases [32].

**Fig 5.**
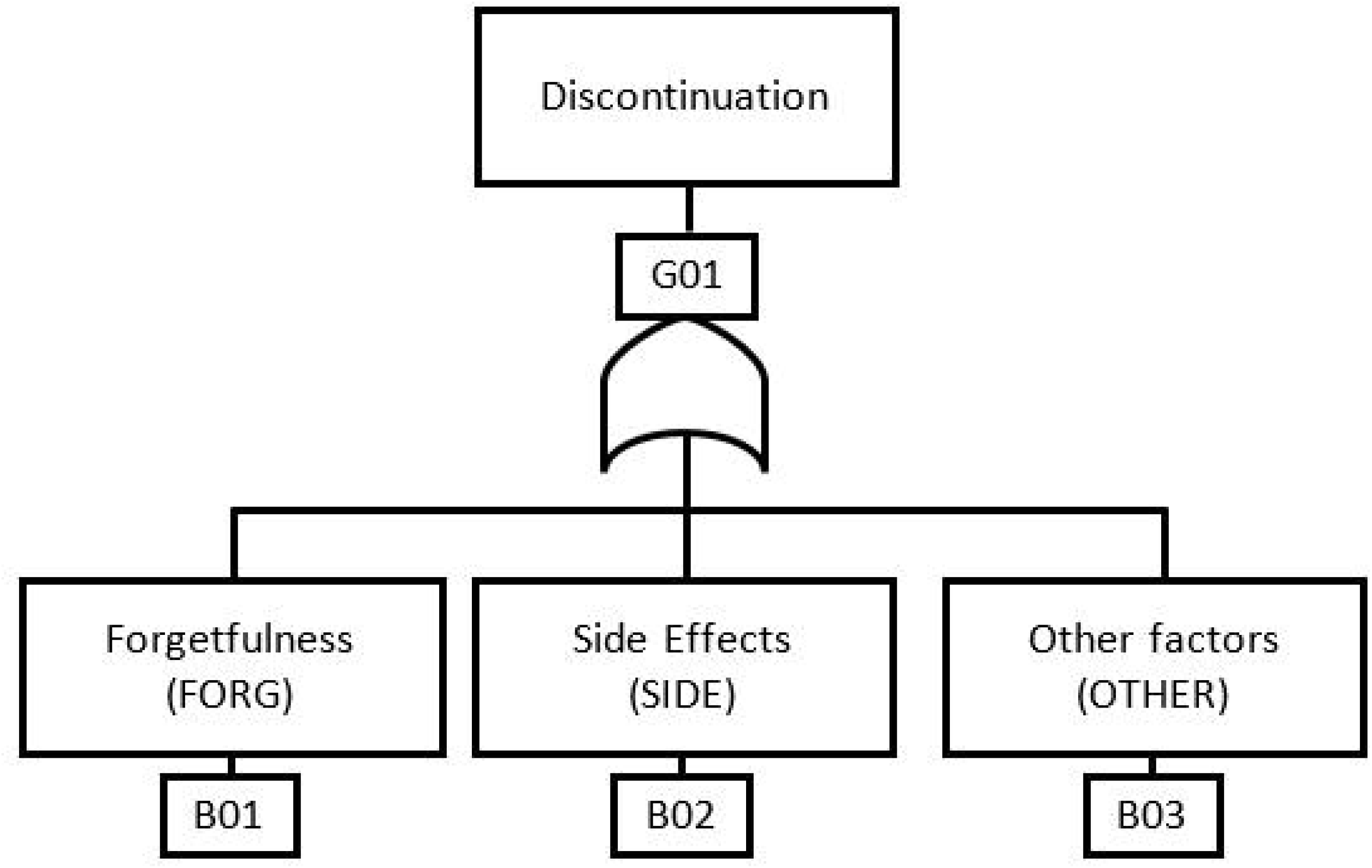
Contribution of NAFs to NA.

This study has established that NATA can reveal the non-adherence factors clinicians need to know to allocate resources targeting those non-adherence factors. It is assumed that, given the information produced by NATA, clinicians decide to reduce *Forgot, Other, NoSym* and *SidEff* by 20% each through measures such as using a pillbox, software app, information/education [33], trust in physician [34] and psychological ownership [35]. The GoldSim model was updated and re-run to determine the impact of the changes on non-adherence; the results are displayed in Fig 6. As expected, the overall non-adherence of the improved system has reduced by nearly 4% at a mean of 0.187, 5% and 95% confident intervals of 0.17 and 0.21 respectively and a standard deviation of 0.39. This reduced the mean number of tablets wasted from 224 to 187 – saving 37 pills that could potentially increase the evaluation of the efficacy of the treatment by 0.37%.

**Fig 6.**
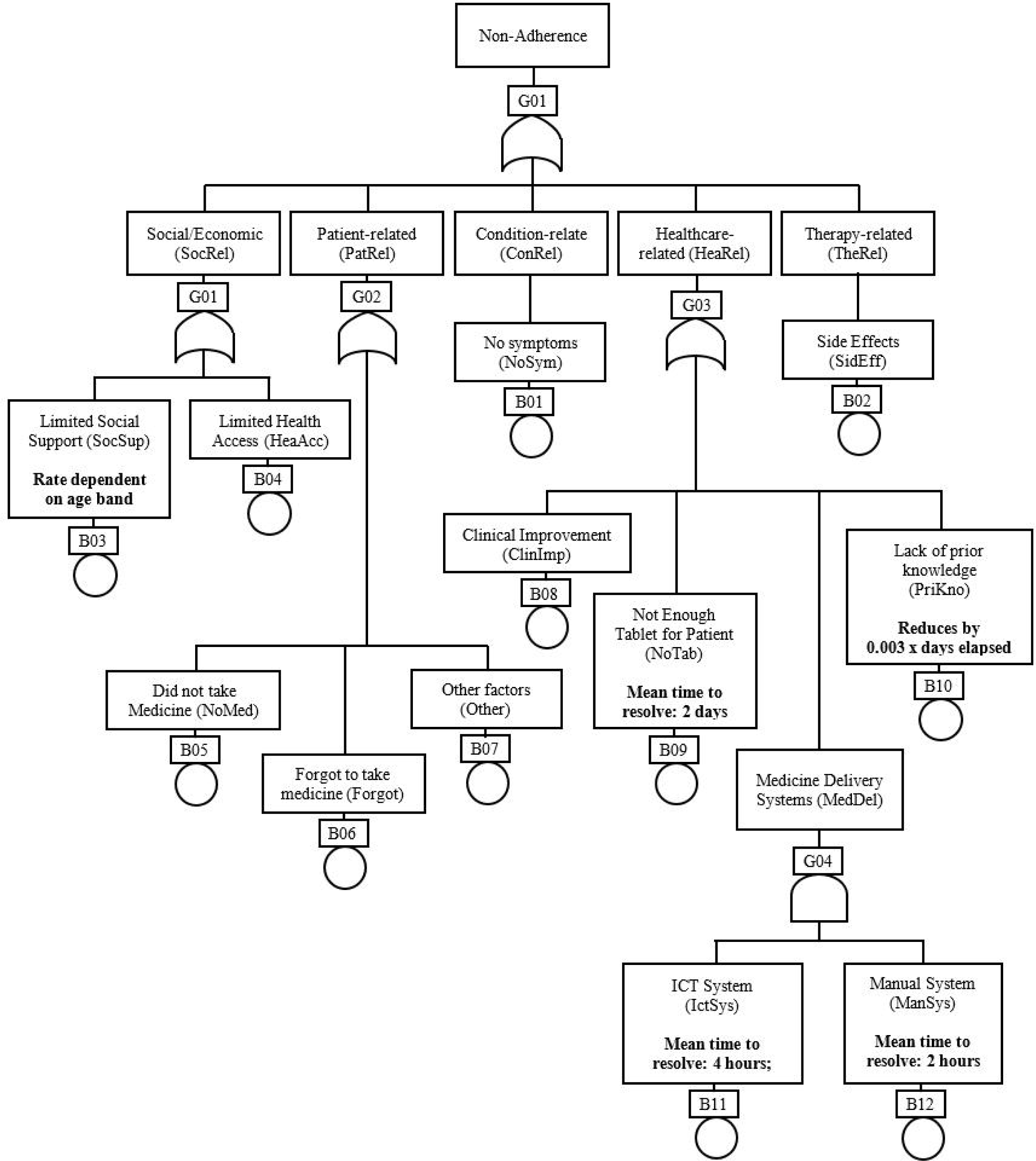
Improvement of Contribution of NAFs to NA.

The significant changes made to the improved model using very generous reduction rates of 20% have enhanced adherence by 3.7% – not as much as one would have expected. The reason for this big change but relatively little impact is that all NAFs would have to be reduced to make significant changes to the overall non-adherence. The results of this case study affirm that no single factor can fully minimise non-adherence [4,11] and provides empirical proof. However, it is still clear that the four main contributing NAFs are *SidEff, Forgot, NoSym* and *Other*; these are factors clinicians should seek to improve to minimise non-adherence.

## Contribution of NATA

Adherence measuring techniques are usually implemented when a treatment has already begun. This study has introduced NATA for predicting patients’ adherence behaviour so that measures can be put in place to improve adherence. NATA is not only a pre-treatment technique; it can also be used during treatment. A NATA model in GoldSim can be updated with changes and re-run to determine how the changes affect the system. This study has proven that NATA can identify non-adherence factors of a treatment regimen and their relationships and contribution to overall non-adherence. With such information, clinicians can implement mitigating strategies to minimise the risk of high non-adherence. Most of the data used in this study – extracted from other studies – are occurrence rates, hence it was assumed they were exponentially distributed. However, the proposed solution, NATA and GoldSim, are not restricted to exponentially distributed rates. Using Monte Carlo simulation, the proposed solution can model and analyse any case study.

## Limitation

This study is not without limitations. The data used in the COVID-19 case study are based on a similar drug – oseltamivir – of a similar disease. The authors assume that the behaviour of COVID-19 patients would be similar to that of the patients who took oseltamivir from six studies. The six studies from which the data was extracted were diverse in terms of demographics and population; therefore, for a geographically specific application, the data may need to be streamlined. The simulation for the case study was modelled to run for ten consecutive days, which is not an accurate reflection of real-world studies where participants of a trial start on different days. NATA is not a stand-alone solution for addressing all the issues with non-adherence; it depends on the results of studies and techniques such as the Medication Adherence Reasons Scale or the Morisky Medication Adherence Scale (MMAS) for data to perform its analysis. In the future, data for NAFs can be sourced from Big Data and/or Artificial Intelligence-enabled systems where possible.

## Conclusion

Non-adherence to a medication regimen is widespread. In addition to financial losses, non-adherence can blur the efficacy of drugs and lead to loss of lives. Most adherence measuring techniques are implemented after the patient has started the medication regimen. This article has explored the use of Fault Tree Analysis (FTA) – an engineering technique for probabilistic risk analysis – to predict the nature of non-adherence. It proposes the Non-Adherence Tree Analysis (NATA) based on classical FTA for modelling and analysing a medication regimen. Based on the results NATA produces, health professionals and clinicians can implement strategies and allocate resources to help improve adherence. NATA can serve as a framework for analysing non-adherence factors in clinical trials and other drug administration processes. The authors have applied NATA to a hypothetical COVID-19 treatment; the results reveal the factors clinicians should concentrate on to minimise non-adherence.

## Data Availability

All data available on request

## Acknowledgements

The authors would like to thank Prof. Peter Naude for reviewing the article and providing constructive comments that have contributed immensely to its quality.

## References

[1] De Geest S, Sabaté E. Adherence to long-term therapies: Evidence for action. Eur J Cardiovasc Nurs 2003;2:323. https://doi.org/10.1016/S1474-5151(03)00091-4.

[2] Wiecek E, Tonin FS, Torres-Robles A, Benrimoj SI, Fernandez-Llimos F, Garcia-Cardenas V. Temporal effectiveness of interventions to improve medication adherence: A network meta-analysis. PLoS One 2019;14:e0213432. https://doi.org/10.1371/journal.pone.0213432.

[3] Osterberg L, Blaschke T. Adherence to medication. N Engl J Med 2005;353:487–97.

[4] Shubber Z, Mills EJ, Nachega JB, Vreeman R, Freitas M, Bock P, et al. Patient-Reported Barriers to Adherence to Antiretroviral Therapy: A Systematic Review and Meta-Analysis. PLOS Med 2016;13:e1002183. https://doi.org/10.1371/journal.pmed.1002183.

[5] ASA, ASCP. Medication Adherence – Where Are We Today □? Overview 2006. http://www.adultmeducation.com/overviewofmedicationadherence_4.html (accessed August 12, 2016).

[6] Vernon A, Fielding K, Savic R, Dodd L, Nahid P. The importance of adherence in tuberculosis treatment clinical trials and its relevance in explanatory and pragmatic trials. PLoS Med 2019;16. https://doi.org/10.1371/journal.pmed.1002884.

[7] Cutler RL, Fernandez-Llimos F, Frommer M, Benrimoj C, Garcia-Cardenas V. Economic impact of medication non-adherence by disease groups: A systematic review. BMJ Open 2018;8:16982. https://doi.org/10.1136/bmjopen-2017-016982.

[8] Wu J-R, Moser DK. Medication Adherence Mediates the Relationship Between Heart Failure Symptoms and Cardiac Event-Free Survival in Patients With Heart Failure. J Cardiovasc Nurs 2018;33:40–6. https://doi.org/10.1097/JCN.0000000000000427.

[9] Costa E, Giardini A, Savin M, Menditto E, Lehane E, Laosa O, et al. Interventional tools to improve medication adherence: Review of literature. Patient Prefer Adherence 2015;9:1303–14. https://doi.org/10.2147/PPA.S87551.

[10] Brown MT, Bussell JK. Medication adherence: WHO cares? Mayo Clin Proc 2011;86:304–14. https://doi.org/10.4065/mcp.2010.0575.

[11] Martin LR, Williams SL, Haskard KB, Dimatteo MR. The challenge of patient adherence. Ther Clin Risk Manag 2005;1:189–99. https://doi.org/10.1089/bar.2012.9960.

[12] Kleinsinger F. The Unmet Challenge of Medication Nonadherence. Perm J 2018;22:18–033. https://doi.org/10.7812/TPP/18-033.

[13] Aldeer M, Javanmard M, Martin RP. A Review of Medication Adherence Monitoring. Appl Syst Innov 2018;1:1–27. https://doi.org/10.3390/asi1020014.

[14] Moon SJ, Lee WY, Hwang JS, Hong YP, Morisky DE. Accuracy of a screening tool for medication adherence: A systematic review and meta-analysis of the Morisky Medication Adherence Scale-8. PLoS One 2017;12. https://doi.org/10.1371/journal.pone.0187139.

[15] Vrijens B, Geest S De, Hughes DA, Przemyslaw K, Demonceau J, Ruppar T, et al. A new taxonomy for describing and defining adherence to medications. Br J Clin Pharmacol 2012;73:691–705. https://doi.org/10.1111/j.1365-2125.2012.04167.x.

[16] Nieuwlaat R, Wilczynski N, Navarro T, Hobson N, Jeffery R, Keepanasseril A, et al. Interventions for enhancing medication adherence. 2014. https://doi.org/10.1002/14651858.CD000011.pub4.

[17] Torres-Robles A, Wiecek E, Cutler R, Drake B, Benrimoj SI, Fernandez-Llimos F, et al. Using dispensing data to evaluate adherence implementation rates in community pharmacy. Front Pharmacol 2019;10:1–9. https://doi.org/10.3389/fphar.2019.00130.

[18] Morisky DE, Ang A, Krousel-Wood M, Ward HJ. Predictive validity of a medication adherence measure in an outpatient setting. J Clin Hypertens 2008;10:348–54. https://doi.org/10.1111/j.1751-7176.2008.07572.x.

[19] Unni EJ, Sternbach N, Goren A. Using the Medication Adherence Reasons Scale (MAR-Scale) to identify the reasons for non-adherence across multiple disease conditions. Patient Prefer Adherence 2019;13:993–1004. https://doi.org/10.2147/PPA.S205359.

[20] Vesely WE, Stamatelatos M, Dugan JB, Fragola J, Minarick J, Railsback J. Fault Tree Handbook with Aerospace Applications. Washington DC: NASA Office of Safety and Mission Assurance; 2002.

[21] Goldsim. Goldsim. A Dyn Simul Approach to Reliab Model Risk Assess Using GoldSim 2020. https://www.goldsim.com/downloads/documents/GoldSim_Reliability_and_PRA.pdf (accessed December 15, 2018).

[22] Brüssow H. The Novel Coronavirus – A Snapshot of Current Knowledge. Microb Biotechnol 2020:1751-7915.13557. https://doi.org/10.1111/1751-7915.13557.

[23] Wang M, Cao R, Zhang L, Yang X, Liu J, Xu M, et al. Remdesivir and chloroquine effectively inhibit the recently emerged novel coronavirus (2019-nCoV) in vitro. Cell Res 2020;30:269–71. https://doi.org/10.1038/s41422-020-0282-0.

[24] Kim J, Combs K, Downs J, Tillman F. Medication adherence: The elephant in the room. US Pharm 2018;43:30–4.

[25] Belmaker I, Lyandres M, Bilenko N, Dukhan L, Mendelson E, Mandelboim M, et al. Adherence with oseltamivir chemoprophylaxis among workers exposed to poultry during avian influenza outbreaks in southern Israel. Int J Infect Dis 2009;13:261–5. https://doi.org/10.1016/j.ijid.2008.06.037.

[26] Chishti T, Oakeshott P. Do general practice patients who are prescribed Tamiflu® actually take it? Br J Gen Pract 2010;60:535. https://doi.org/10.3399/bjgp10X514891.

[27] Choo D, Hossain M, Liew P, Chowdhury S, Tan J. Side effects of oseltamivir in end-stage renal failure patients. Nephrol Dial Transplant 2011;26:2339–44. https://doi.org/10.1093/ndt/gfq737.

[28] McVernon J, Mason K, Petrony S, Nathan P, LaMontagne AD, Bentley R, et al. Recommendations for and compliance with social restrictions during implementation of school closures in the early phase of the influenza A (H1N1) 2009 outbreak in Melbourne, Australia. BMC Infect Dis 2011;11:1–7. https://doi.org/10.1186/1471-2334-11-257.

[29] Strong M, Burrows J, Stedman E, Redgrave P. Adverse drug effects following oseltamivir mass treatment and prophylaxis in a school outbreak of 2009 pandemic influenza a(H1N1) in June 2009, Sheffield, United Kingdom. Eurosurveillance 2010;15:1–6. https://doi.org/10.2807/ese.15.19.19565-en.

[30] Wallensten A, Oliver I, Lewis D, Harrison S. Compliance and side effects of prophylactic oseltamivir treatment in a school in South West England. Eurosurveillance 2009;14:1–4. https://doi.org/10.2807/ese.14.30.19285-en.

[31] Gu L, Wu S, Zhao S, Zhou H, Zhang S, Gao M, et al. Association of social support and medication adherence in Chinese patients with type 2 diabetes mellitus. Int J Environ Res Public Health 2017;14:1522. https://doi.org/10.3390/ijerph14121522.

[32] Nishiura H, Kobayashi T, Suzuki A, Jung S-M, Hayashi K, Kinoshita R, et al. Estimation of the asymptomatic ratio of novel coronavirus infections (COVID-19). Int J Infect Dis 2020. https://doi.org/10.1016/j.ijid.2020.03.020.

[33] Agyepong IA, Ansah E, Gyapong M, Adjei S, Barnish G, Evans D. Strategies to improve adherence to recommended chloroquine treatment regimes: A quasi-experiment in the context of integrated primary health care delivery in Ghana. Soc Sci Med 2002;55:2215–26. https://doi.org/10.1016/S0277-9536(01)00366-5.

[34] Lee YY, Lin JL. The effects of trust in physician on self-efficacy, adherence and diabetes outcomes. Soc Sci Med 2009;68:1060–8. https://doi.org/10.1016/j.socscimed.2008.12.033.

[35] Mifsud M, Molines M, Cases AS, N’Goala G. It’s MY health care program: Enhancing patient adherence through psychological ownership. Soc Sci Med 2019;232:307–15. https://doi.org/10.1016/j.socscimed.2019.05.015.

